# Development of an index to assess Covid-19 hospital care installed capacity in the 450 Brazilian Health Regions

**DOI:** 10.1101/2021.06.17.21259121

**Authors:** Claudia Cristina de Aguiar Pereira, Fernando Ramalho Gameleira Soares, Gustavo Saraiva Frio, Carla Jorge Machado, Layana Costa Alves, Fernando José Herkrath, Rodrigo Tobias de Sousa Lima, Ivana Cristina de Holanda Cunha Barreto, Everton Nunes da Silva, Anny Beatriz Costa Antony de Andrade, Leonor Maria Pacheco Santos

## Abstract

**Objective:** We assessed the Brazilian health system’s ability to respond to the challenges imposed by the Covid-19 pandemic considering hospital capacity in the 450 Health Regions of the country in 2020. Hospital capacity referred to the availability of hospital beds, equipment, and human resources.

**Methods:** Data came from National Register of Health Facilities on the availability of Covid-19 resources in inpatient facilities from January to December,2020. Assessed resources were health professionals, hospital beds, and medical equipment. A synthetic indicator, Installed Capacity Index (ICI) was calculated using Principal Component Analysis.

**Results:** There was an increase in all selected indicators between January and December 2020. We observed differences between the Northeast, North regions, and the other regions of the country. Most Health Regions presented low ICI. The ICI increased especially in regions with considerably high baseline capacity in January 2020. The Northeast and North had a higher concentration oflow ICI regions.

**Conclusion:** The information here provided may be used by health authorities, providers, and managers in planning and adjusting for future Covid-19 care and in dimensioning the adequate supply of hospital beds, health care professionals, and devices in Health Regions to reduce associated morbidity and mortality.

## INTRODUCTION

The Covid-19 pandemic has challenged health systems worldwide^1^ and in Brazil where a public (Sistema Unico de Saude - SUS) and private health systems coexist. To face the pandemic, health services must be structured and able to adapt to emerging needs. Thus, in the context of treatment for those affected by the disease, actions are needed to plan and assess the installed capacity of health services to provide timely and adequate hospital care in different country regions.

The provision of health services depends on the installed capacity of health facilities to provide health care in due time and place, especially in facing the Covid-19 pandemic. Adequate hospital treatment can provide satisfactory outcomes for patients with Covid-19^2^, but this demands, especially in severe cases that require hospitalization in Intensive Care Unit (ICU), specific equipment, specialized and trained human resources, and the provision of enough beds to meet the demand at any point in time^3-5^. In Brazil, what has been observed in the first year of the pandemic in several circumstances is that supply did not match demand, even though there has been an increase in hospital beds^6^. The uneven distribution among the country regions, with a deficit of beds in most Brazilian municipalities^7^ mainly in the North and Northeast regions, has been a significant factor in the loss of lives^6^. It is important to describe the hospital capacity to care for Covid-19 cases, thinking about the most critical supply elements, namely beds, devices and human resources^4,8,9^, understanding its evolution over time since the beginning of the pandemic.

This study aims to measure the installed capacity of all Brazilian hospitals (public and private) to care for Covid-19 cases in the country’s official Health Regions in 2020. Installed capacity refers to the availability of hospital beds, devices, and human resources.

## METHODS

This is a longitudinal analytical study carried out with data from the National Register of Health Facilities (CNES) regarding the availability of health professionals, hospital beds, and medical equipment in all hospitals registered in the system, both public and private, during the period from January to December 2020, distributed in the officially established 450 Health Regions in Brazil. A Health Region comprises a group of municipalities that share their resources, such as hospitals, emergency units, and primary care and financial resources into care networks for the provision of services to the populations of the municipalities. The regionalization of the Unified Health System (SUS) was determined as a strategy to optimize the management of the system, the rationalization of resources, and the institutional contribution to creating health care networks ^10^. Public domain data made available by the Ministry of Health were used, whose last update was on 01/15/2021.

Regarding the devices or equipment, computed tomography (CT scanners), defibrillators, ECG monitors, ventilators, and resuscitators were included in the analysis, as they are considered necessary in treating severe cases of COVID-19^5^. All the devices accounted for had an indication in the CNES as “in use”. For hospital beds, only those types used in caring for patients with Covid-19 were taken into account. Two categories were proposed: the first with hospital beds of the clinical type, intermediate care, and ventilator support; the second with adult ICU beds in general or exclusive to Covid-19.

In accounting for health professionals, we considered the staff linked to hospitals that had at least one of the bed categories proposed. Among the professional specialties that work in these facilities, only certified nursing assistants, nurses, physicians, and physical therapists were used. This specification is justified by the more direct action of these workers in intensive care of patients diagnosed with Covid-19, a decision that is in line with the requirement of specialists for the qualification and implantation of an essential team in ICUs exclusive or not for patients with Covid-19 (MB/Ordinance No. 3,432, OF AUGUST 12^th^, 1998).

The number of hospital beds, devices, and health professionals in use or available in health facilities was broken down according to the type of funding or availability of equipment/beds/professionals. The supply of hospitals was stratified into two categories: those belonging exclusively to SUS (public system) and beneficiaries of private health insurance plans and/or private users. Although registered individually with their connection to the hospitals, the records were accounted for on the scale of the municipality and the (micro) Health Region where the hospital was located. Rates of beds, devices, and health professional availability were obtained, adding SUS and private (total) and exclusively in SUS (public only). To calculate the total rates, population data were obtained from the population estimates provided by the Brazilian Institute of Geography and Statistics (IBGE), with a reference date of July l^st^, 2020. The non-SUS data considered the number of beneficiaries of health insurance plans for medical assistance that included dental assistance or not, made available at the municipal level every quarter by the National Health Agency (ANS) in March, June, and September 2020, with municipal distribution of the number of beneficiaries. The total number of SUS users was also considered by subtracting health insurance plan beneficiaries from the total municipal population.

To obtain the data, the *Microdatasus* library^11^ developed in R language was used, which allowed downloading and pre-processing the data available in the DATASUS platform to be carried out as a batch.

Initially, means and measures of relative dispersion of regional values around the mean were calculated, comparing total and exclusive SUS availability. We sought to represent a month before the pandemic and another during it. Therefore, we chose the months of January and December 2020, respectively.

We calculated total rates per 10,000 inhabitants for the number of devices, beds, and professionals in public and private networks. SUS rates were restricted to devices and professionals available exclusively in the Unified Health System, for which the population considered in the denominator were individuals who do not have private health insurance plans. In the construction of the index, the Principal Component Analysis (PCA) method was used to present an empirical summary of the original dataset. The use of PCA allows the explanation of the variance and covariance structure of a vector of random variables by constructing linear combinations of the original variables^12^. PCA can reduce a large number of variables to a model composed of *k* main components that are not correlated with each other and can explain a relevant proportion of the total variance of the original variables. The existence of multicollinearity between the original variables is not, unlike in other techniques, a problem for calculating the component. We sought to obtain an indicator that captured the common behavior and, in a simplified way, from its linear convergence, observed in the original variables between the Health Regions, in addition to the time variation of these values.

Following guidelines from Mingoti ^12^, to avoid excessive influence of discrepancies in the variances of the original variables, we chose to standardize the variables by their mean and standard deviation, where each of the 11 original variables *X*_*i*_ were transformed, obtaining *Z*_*i*_ = (*X*_*i*_ − *μ*_*i*_)/*σ*_*i*_, where *E*(*X*_*i*_) = *μ*_*i*_ and *Var*(*X*_*i*_) = *σ*_*i*_^2^, with *i* = 1,2, …, *n* = 11. At the end of the analysis, each of the resulting *k* main components could be calculated from simple linear equations with multiplicative constants *β*_*j,i*_ (*j* = 1,2, …, *k*) associated with each variable and identified by standardized eigenvectors for each component (ANNEX 1).

The statistical software used was SPSS 26.0. The PCA was applied to monthly data from all Health Regions as separate records to allow for a time analysis, from which the indicator of the first component generated was obtained. The indicator synthesized most of the data variance, an expected result, given the relevant correlations between the original variables.

The calculation of the indicator allowed exploring regional differences in the installed capacity of hospital services in offering adequate care to patients affected by Covid-19, who presented severe conditions of the disease and needed hospital care and hospitalization.

To assess whether a principal component analysis could generate a model adequately adjusted to the original data, two adequacy measures were used: the Kaiser-Meyer-Olkin criterion (KMO) and Bartlett’s test of sphericity^12^ first test, an adequate adjustment is considered when KMO > 0.8 is obtained. With Bartlett’s test, which consists of a chi-square test that assesses the null hypothesis that the correlation matrix between the original variables is equal to the identity matrix, an adequate adjustment is reached in the rejection of the null hypothesis with a p-value of at least 0.05.

In order to facilitate the analysis of essential aspects of the ICI, an interpretative strategy was chosen to stratify the index into groups of value ranges where differences in the behavior of Health Regions were verified at different levels of installed capacity. The intervals for the groups were defined by applying the Jenks algorithm (“natural breaks”), which is an algorithm commonly implemented in geographic information systems (GIS) to stratify a data series where each class tends to have a smaller internal variance and at the same time presents a greater variance between different classes.

## RESULTS

### Descriptive Statistics of Original Variables

When comparing the total and exclusively SUS rates, on average, it is possible to notice a more relative supply of resuscitators, respirators/ventilators, nursing professionals, physical therapy, physicians, and certified nursing assistants in SUS than in the total group, which also incorporates the private supply. The total rates indicate an advantage of the SUS only for the relative presence of defibrillators, ECG monitors, CT scanners, and hospital beds (either clinical or ICU).

It should be noted that there was an increase in all indicators between January and December 2020, both in Total and exclusively SUS. There was an increase of 192.9% in the rate of hospital beds in adult ICU in the Total.

The data in Table 1 for the total rates in December 2020 are presented by Health Region (Figure 1). The values were standardized - the calculation is the ratio between the value of the variable minus the mean and the standard deviation. The variables are divided into five groups in which shades of red represent the groups with the lowest proportion and shades of gray and black represent the highest proportion. The North and Northeast regions presented few Health Regions in shades of gray and black (when considering all variables) and many regions in shades of red, mainly in equipment. The regions with the highest proportion of devices, hospital beds, and health professionals are the South and Southeast.

**Table 1.**
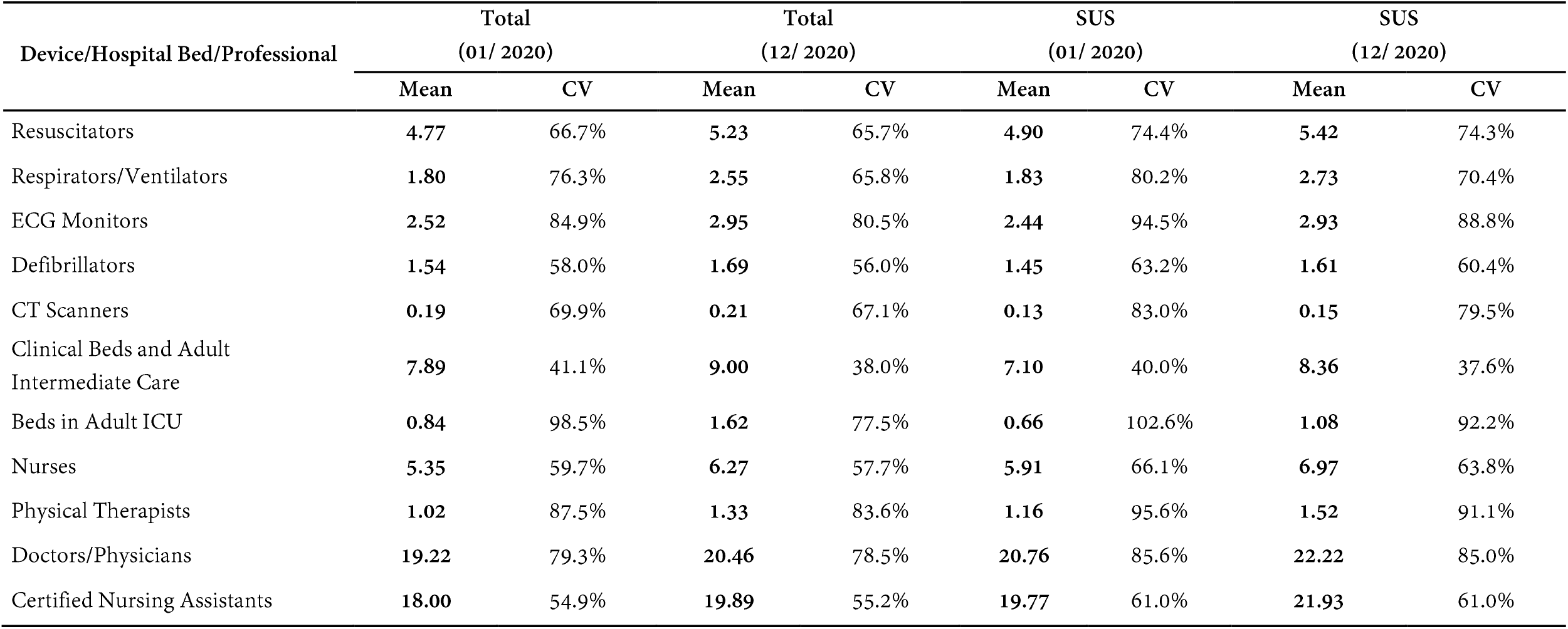
Rates of devices, hospital beds, and health professionals per 10,000 inhabitants/users coefficient of variation and mean values, according to total or SUS-only availability. Brazilian Health Regions, January, and December 2020.

**Table 2.** Communalities of the Principal Component Analysis (PCA) and loading of the original variable in relation to the first component. Brazilian Health Regions, 2020.

| Device /Hospital Bed/Professional | Total |  | SUS |  |
| --- | --- | --- | --- | --- |
|  | Communalities PCA Total | Loading Component 1 | Communality PCA SUS | Loading Component 1 |
| Resuscitator | 0.706 | 0.840 | 0.679 | 0.824 |
| Respirator/Ventilator | 0.847 | 0.920 | 0.836 | 0.914 |
| ECG Monitor | 0.808 | 0.899 | 0.775 | 0.880 |
| Defibrillator | 0.716 | 0.846 | 0.680 | 0.825 |
| CT Scanner | 0.462 | 0.680 | 0.381 | 0.617 |
| Clinical Beds and Adult Intermediate Care | 0.229 | 0.478 | 0.223 | 0.473 |
| Beds in Adult ICU | 0.753 | 0.868 | 0.719 | 0.848 |
| Nursing Professionals | 0.701 | 0.837 | 0.726 | 0.852 |
| Physical Therapy Professionals | 0.657 | 0.810 | 0.664 | 0.815 |
| Medical Professionals | 0.742 | 0.861 | 0.792 | 0.890 |
| Certified Nursing Assistant Professionals | 0.760 | 0.872 | 0.798 | 0.893 |

**FIGURE 1.**
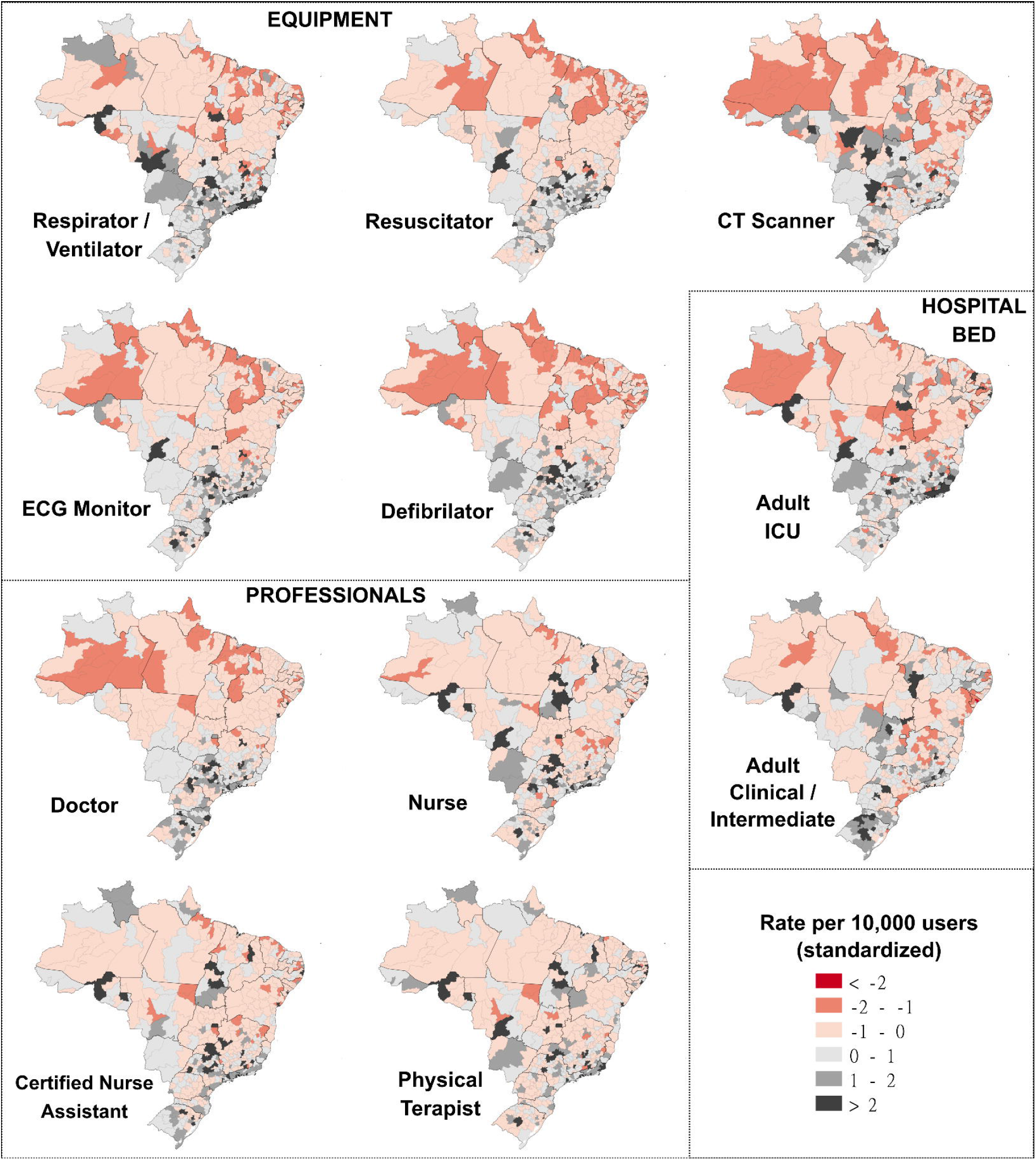
Standardized rates of Equipment, Professionals and Hospital Beds per 10,000 users.

### Principal Component Analysis Results

The use of the KMO test for the 22 original variables, 11 Total and 11 SUS, performed separately, generated high KMO indexes, with values of 0.925 and 0.930, respectively, therefore, above the recommendation for an adequate adjustment (KMO > 0.8). Bartlett’s test showed results that satisfactorily made it possible to reject the test’s null hypothesis for both PCAs (p<0.05).

The PCAs, with both total and exclusive data from SUS, resulted in only one component, each with an eigenvalue greater than 1, which in the literature is a sufficient criterion for choosing only this component to explain the original variables. For total data, component 1 can explain 67.1% of the data variance, while component 1 of the PCA for SUS only data can explain 66.1% of the variance of the original data.

Communality represents the proportion of variance for each variable included in the analysis explained by the extracted components. The component loadings indicate the degree of correlation between the original variable and the component. Both loadings and commonalities have a higher value in respirator/ventilator and a smaller one in clinical beds and adult intermediate care.

### Installed Capacity Index (ICI)

To meet the practical objective of proposing an indicator capable of capturing regional differences and the evolution of installed hospital capacity in coping with Covid-19, added to the intention of synthesizing both public and private capacity into a single indicator, we chose to present only a single index of installed capacity that derives from the component obtained in the PCA with the total data.

The use of the first principal component as a synthetic indicator was found to be sufficiently representative, thus here proposed as the Installed Capacity Index (ICI). Considering the vector *Z* = [*Z*_1_, *Z*_2_, …, *Z*_11_] of the standardized total rates, or even rewriting the equation in the function of the original variables, the ICI is obtained from the explicit equations in Annex 1. Keeping in mind the limitation that the index uses only data from 2020 as parameters, it is possible to calculate, for comparison purposes, the index of each region for any month of competence, including for more recent periods.

Figure 2 shows the evolution of the ICI between January and December 2020, indicating growth in all regions of Brazil, increasing the number of Health Regions in groups 4 and 5 of installed capacity (higher capacity). The North and Northeast regions have most of their Health Regions belonging to Group **1** and 2, showing lower ICI.

**FIGURE 2.**
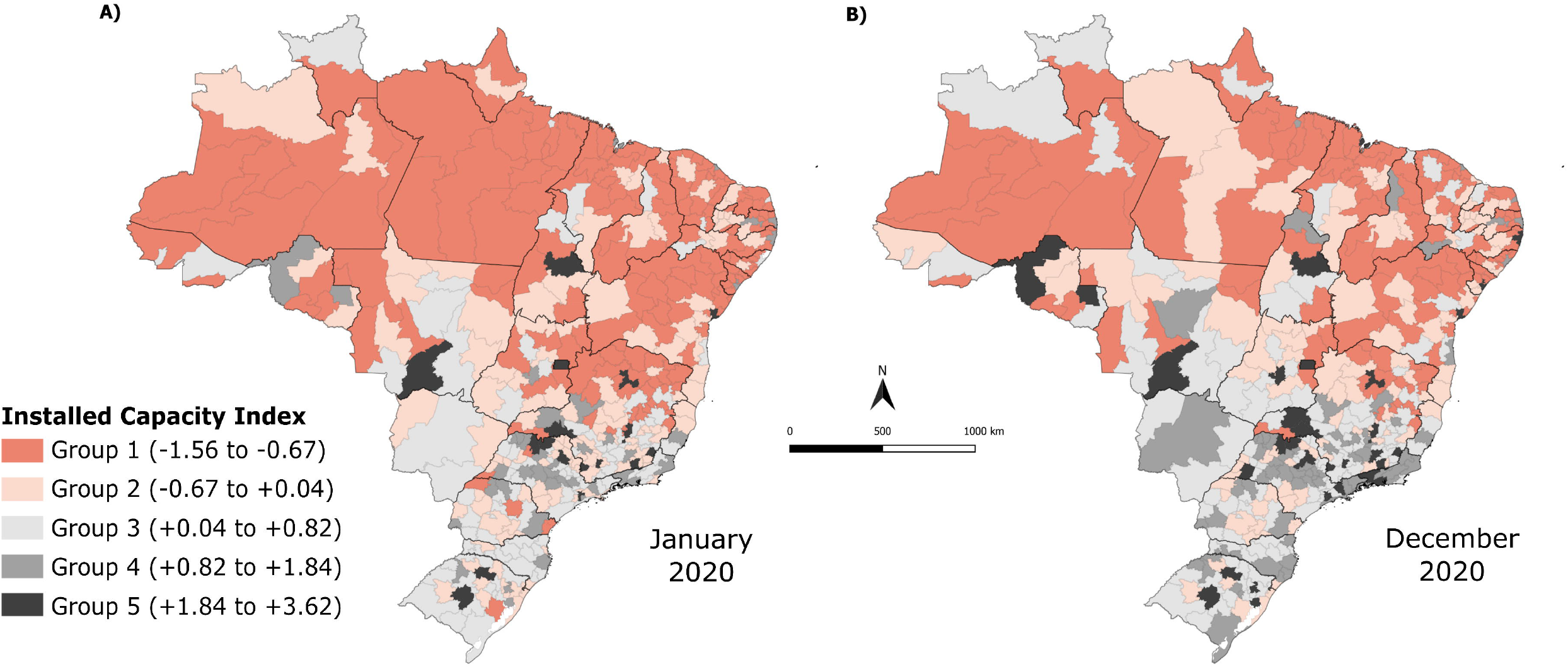
Installed Capacity Index in Brazilian Health Regions, January and December, 2020.

Figure 3 is stratified by groups according to the group where the Health Region was in December 2020. There is an evident increase in ICI in most groups, except for Group 1, which corresponds to the group with the lowest ICI.

**FIGURE 3.**
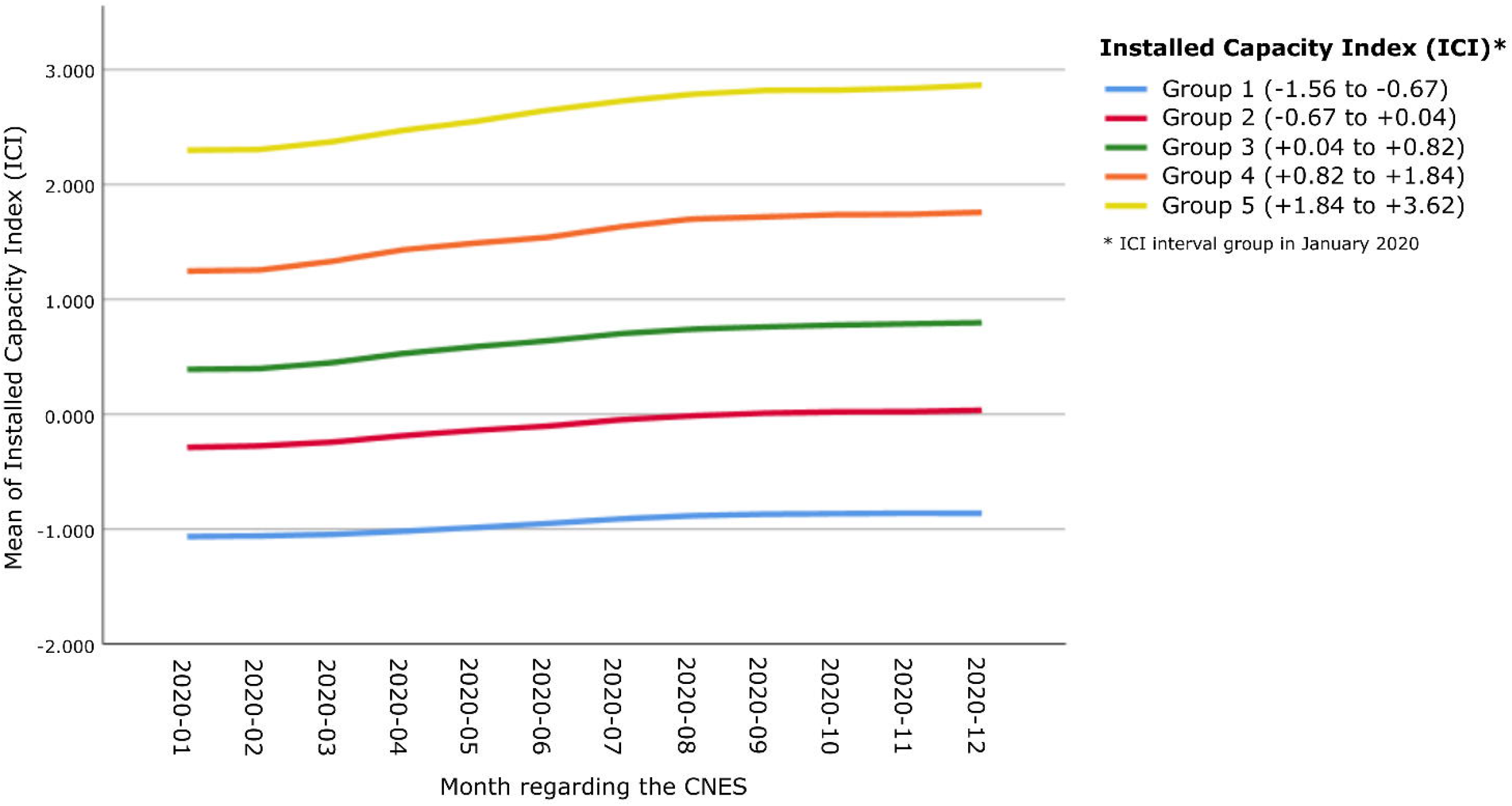
Mean Installed Capacity Index by month, 2020.

The histograms of the difference from the end of 2020 to the beginning of the year observed by groups of ICI intervals in January 2020 (Figure 4) show an exceedingly small number of Health Regions that presented decrease in their installed capacity (negative difference) and a growth between 0 and 0.5 points in the index in most regions. The ICI has a positive, albeit weak (*R*^2^ adjusted of 0.252) correlation, with the differences observed from January to December in relation to the index itself, to certain extent corroborating the tendency of higher ICI growth in Health Regions that already had higher ICI values in January 2020.

**FIGURE 4.**
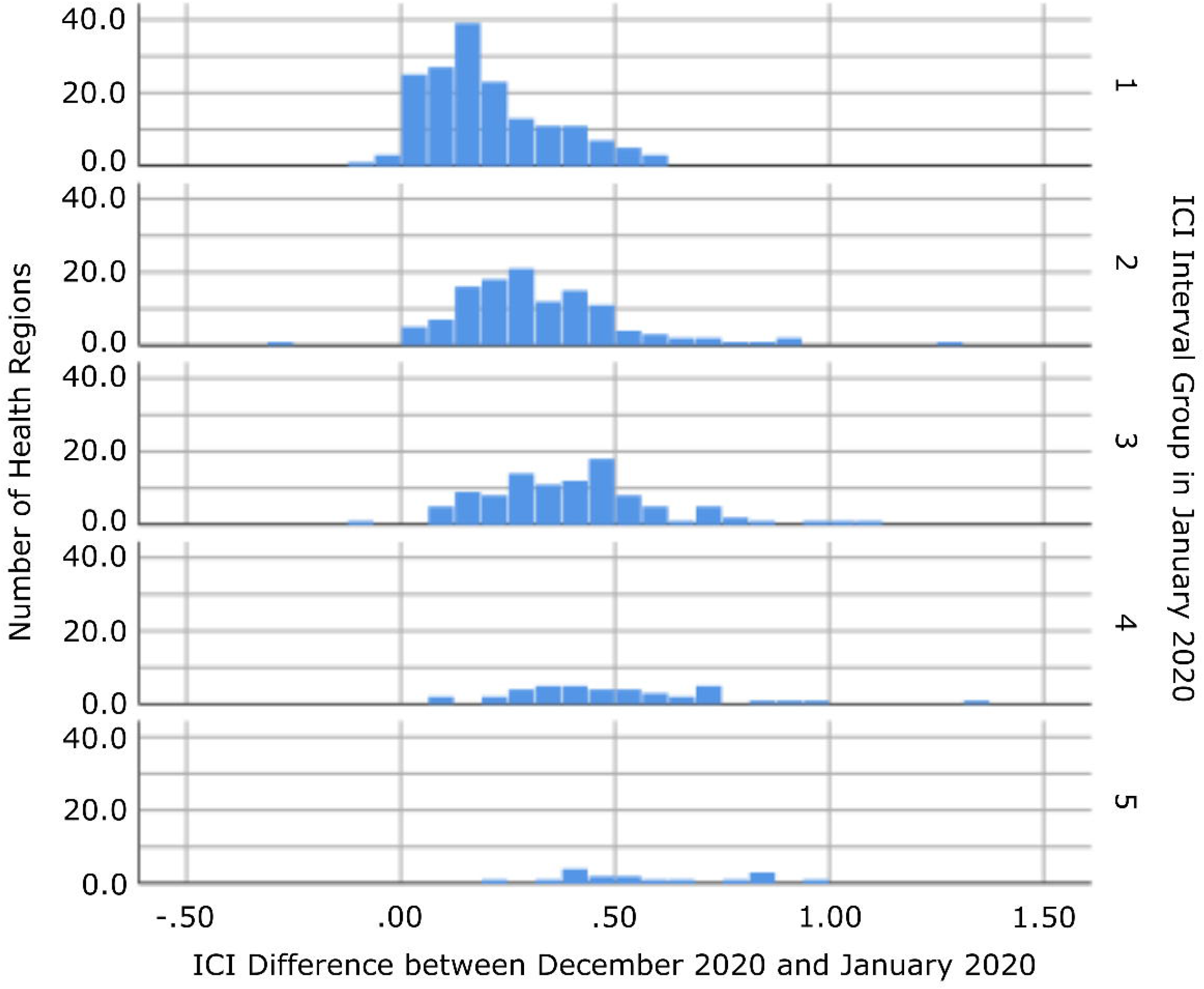
Difference in Installed Capacity Index between December and January, 2020.

## DISCUSSION

The present study is the first to present an installed capacity index (ICI) for hospital supply of care for Covid-19 patients, considering the country’s official Health Regions. Its importance lies in the fact that the Covid-19 pandemic made it essential to measure the capacity to provide the necessary health care, monitor and measure any expansions in the response over the months of the pandemic.

First, there was an increase in all selected indicators between January and December 2020, both for the total and SUS-exclusive rates. Second, it was possible to observe differences between the Northeast and North regions and the other regions of the country, in which the majority of Health Regions presented low ICI. Third, the evolution of the ICI was observed between the beginning and the end of 2020, with an increase in the number of Health Regions in the groups with greater installed capacity, although the average increase in the ICI was more evident in the groups that already had high capacity in January 2020 (baseline).

The findings about Health Regions belonging to the Northeast and North regions are concerning, as they show the absence of improvements towards the disparities already observed in studies with cross-sectional data from CNES referring to the period before the pandemic^13,5,4,14^.Moreira ^14^ pointed out that in January 2020, almost 45% of the Health Regions that did not have an ICU were located in the Northeast region. Another study identified that the state of Amapa did not register any municipality on the list of those with initial structural capacity in February 2020 to treat severe Covid-19 cases, considering the availability of ICU beds and other devices listed as necessary. At the same time, Sergipe, Amazonas, and Roraima states presented only one municipality on the list ^45^there were also significant care gaps regarding the availability of beds in the North, Northeast, and Midwest regions, emphasizing the absence ofICU beds in more than a quarter of the Health Regions in February 2020. The study carried out by Bezerra et. al.^15^, with CNES data from April 2020, when the pandemic was already underway, to calculate an infrastructure index for the states or Federation Units (UF), showed that the UF with the worst indexes were Amapa and Roraima (located in the North) and the best ranked were Sao Paulo and Minas Gerais (Southeast region). On the other hand, it is best to focus on more specific territorial sections such as Health Regions (macro or micro-regions) or municipalities since more aggregated analyses may fail to capture important differences within the UF. Noronha et. al.^3^ showed a worse situation for macro and micro Health regions in the North and Northeast regions concerning the availability of hospital beds and assisted ventilation equipment, indicating a greater probability of system collapse even with low infection rates.

There was an advantage of the total supply (SUS and private) compared to SUS alone for the presence of defibrillators, ECG monitors, CT scanners, and hospital beds (either clinical or ICU). This finding is relevant, as it shows an expanded supply of clinical beds and ICU when considering the two health systems. Supplying hospital beds is of paramount importance in the treatment of moderate and severe cases of Covid-19. Requia et. al.^7^ estimated the deficit in the capacity of hospital beds in the municipalities of Brazil at the beginning of the pandemic, showing a worse scenario in the municipalities of the North and Northeast macro-regions. The study also suggested that restrictions on social contacts and an increase in hospital capacity could promote an improvement in this scenario^7^. In this regard, the present study observed an increase of almost 193% in the total rate of adult ICU beds between January and December 2020, although it did not occur equally among the Health Regions of the country. Conte et. al.^6^ also showed that the expansion of ICU beds did not reduce the inequities between regions and was not able to solve the insufficiency of hospital beds before the Covid-19 pandemic nor the provision of more health human resources.

An analysis of the first 250,000 hospitalizations in Brazil showed higher mortality in the North and Northeast regions. It also showed lower rates of hospital beds and ICUs in these regions and the concentration of these resources in state capitals. The difference in hospital mortality is also a consequence of inequities in the availability and access to services, reflecting the frailty of the health system in these regions.^16^ The higher mortality in these regions had already been pointed out in the first months of the pandemic.^17^ The regional inequalities observed in installed capacity and the greater difficulty in increasing the number of Health Regions with capacity shows the historically existing difficulties in the regionalization of health in Brazil and the low response capacity of the system to modify this scenario in a short period in the face of need.^6,18^

The problem of installed capacity for hospital care, exposed by the present study, refers to three structural issues related to the funding of the health system, national autonomy in technological development and the production of devices for the service network, and the training and provision of health professionals.

Regarding the first issue, it is known that SUS, a health system whose principles are universally inscribed, is the only one in the world to provide care to more than 100 million people. However, what is enshrined in the Constitution and the Organic Health Laws has never been made possible because of insufficient funding. To make matters worse, in 2016, Amendment No. 95 was approved, which froze public spending in health for 20 years. The context of increased poverty in the country and an aging population, associated with inflation in the health sector, has caused severe underfunding. ^19^

The second issue is related to the technological dependence from other countries, thus resulting in high levels of importation of medical devices and other health items. Brazilian researchers have been investigating and advocating for the development of the Health Economic-Industrial Complex for years, understanding that the construction of a health system that allows the constitutional precepts of universality and wholeness^20^ to be realized must be associated with the consolidation of a productive base and innovation in health so that the national health system is structurally sustained. In a continental country like Brazil, with more than 200 million people, the existence of a productive health system, involving the industry and health services in an articulated way, presents itself as a condition without which access to health cannot be guaranteed struct urally.^21^ The dependence on other countries to provide devices for expanding the number of ICU beds left the Brazilian population and health workers very vulnerable during the pandemic in 2020 and points to the need to face and overcome them.^18^

The third issue is related to the training of human resources for health, especially doctors, nurses, certified nursing assistants, and specialized physical therapists, hospital care, and intensive care, in sufficient numbers to guarantee universal and comprehensive healthcare^22^. which requires medium and long-term planning, especially for the training of specialists and their equitable provision in all Health Regions. In the context of the outbreak of Covid-19 cases, this shortage of professionals is more evident. ^23^

It is worth emphasizing the importance of using CNES as a rich data source on the availability of hospital beds, equipment, and health professionals. In this database, all health establishments, public and private, of individuals or legal entities are registered. Comprehensively and nationally, it is the only public and unrestricted database capable of providing updated data, thus being of great value for decision making. The proposal for an index encompassing several aspects of installed capacity is based on the principle that, in addition to the expansion of beds, the availability of devices and professionals is necessary to assess an adequate response capacity of the health system.^24^ Nevertheless, investments should not be restricted to the provision of intensive care since other measures in public health, such as the articulation between epidemiological surveillance and primary health care, physical distancing, testing, contact tracing, and health communication, could be more efficient in saving lives in the short term without vaccine availability in 2020 to prevent individuals from getting sick in the first place.^25,26^

It is vital to present some limitations identified in the present work. Despite the positive aspects of CNES as a data source, this system depends on self-completion and updating by the facilities, and for this reason, it can lead to errors or inaccuracies. Therefore, the study is limited to aspects of the reported supply of resources, and it does not reach the dimension of the demand for the use of these resources in the different Health Regions of the country. However, these difficulties sought to be overcome, given that the study used a relative measure to allow comparability of installed capacity between Health Regions.

## CONCLUSION

The present study’s findings allowed the understating of inequalities in the installed capacity to care for patients affected by Covid-19 in the Health Regions of Brazil. We identified a concentration of regions with lower values of the capacity index in the Northeast and North regions of the country and inequality in the increase in the indicator over time, despite the average increase observed. These findings show the need to expand the availability of hospital beds, professionals, and devices in these regions to better cope with Covid-19 and to decrease associated morbidity and mortality. Future monthly follow-up with the ICI presented here will allow monitoring the supply dynamics and guide adjustments in the service supply and improvements in the organization of the care networks in the Health Regions of the country.

## Data Availability

Article used public and free data.

## ABBREVIATION LIST

ANS: Brazilian National Health Agency
CNES: National Register of Health Facilities
ECG: Electrocardiogram
IBGE: Brazilian Institute of Geography and Statistics
ICI: Installed Capacity Index
ICU: Intensive Care Unit
KMO: Kaiser-Meyer-Olkin criterion
PCA: Principal Component Analysis
SUS: Sistema Dnico de Sau.de (Brazilian Public Health System)

## Expanded Method

### Annex 1

- *Score Equation of Principal Components* Given the eigenvector 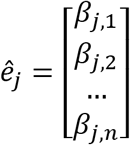, the following equation is obtained:

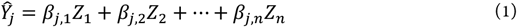 *Ŷ*_*j*_ *= score of component j resulting from the analysis of principal components* *β*_*j,k*_ *= multiplicative constant of the k-th original standardized variable associated with component j* *Z*_*k*_ *= value of the k-th original standardized variable for a given health region*
- *Complete Equation of Installed Capacity Index (ICI) with standardized variables*

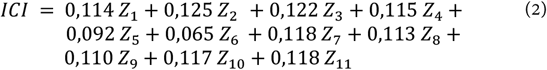 Where: *Z*_1_ *= standardized value of resuscitators per 10 thousand users* *Z*_2_ *= standardized value of respirators/ventilators per 10 thousand users* *Z*_3_ = *standardized value of ECG monitors per 10 thousand users* *Z*_4_ *= standardized value of defibrillators per 10 thousand users* *Z*_5_ = *standardized value of CT scanners per 10 thousand users* *Z*_6_ *= standardized value of clinical beds/intermediate care per 10 thousand users* *Z*_7_ = *standardized value of lCU beds per 10 thousand users* *Z*_8_ *= standardized value of nurses per 10 thousand users* *Z*_9_ = *standardized value of physical therapists per 10 thousand users* *Z*_10_ **=** *standardized value of doctors per 10 thousand users* *Z*_11_ **=** *standardized value of certified nursing assistants per 10 thousand users*
- *Derivation of the Complete Equation of the Installed Capacity Index (ICI) as a function of the original variables*

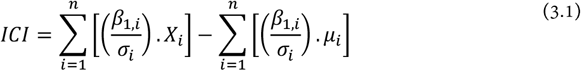

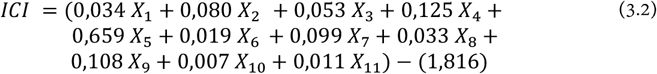 Where: *X*_1_ *= original value of resuscitators per 10 thousand users* *X*_2_ *= original value of respirators/ventilators per 10 thousand users* *X*_3_ *= original value of ECG monitors per 10 thousand users* *X*_4_ *= original value of defibrillators per 10 thousand users* *X*_5_ *= original value of CT scanners per 10 thousand users* *X*_6_ *= original value of clinical beds/intermediate care per 10 thousand users* *X*_7_ *= original value of lCU beds per 10 thousand users* *X*_8_ *= original value of nurses per 10 thousand users* *X*_9_ *= original value of physical therapists per 10 thousand users* *X*_10_ *= original value of doctors per 10 thousand users* *X*_11_ = *original value of certified nursing assistants per 10 thousand users*

